# EFFECTS OF MODIFIED PROGRESSIVE MUSCLE RELAXATION COMBINED WITH SLOW STROKE BACK MASSAGE ON BLOOD PRESSURE REDUCTION, STRESS ALLEVIATION, AND ENHANCED SLEEP QUALITY IN OLDER ADULTS: A LITERATUR REVIEW

**DOI:** 10.1101/2025.01.05.25320000

**Authors:** Asriadi, Moses Glorino Rumambo Pandin, Nursalam

## Abstract

**Background:** Progressive muscle relaxation is a systematic technique used to achieve deep states of relaxation and has been proven to improve the quality of life related to both medical and non-medical conditions. The goal is to consciously experience the difference between tension and relaxation, just like applying pressure to the muscles, where all the muscles in the body from the tip of the head to the tip of the toes, which were initially tense, become relaxed after receiving progressive pressure. Therefore, an explanation is needed that discusses the influence of modified slow stroke back massage progressive muscle relaxation on reducing blood pressure, stress, and sleep quality in the elderly.

**Objective:** to identify the effects of progressive muscle relaxation and slow stroke back massage on the reduction of blood pressure, stress, and sleep quality in the elderly.

**Method:** a scope review of this research by searching for articles related to the effects of progressive muscle relaxation and slow stroke back massage on the reduction of blood pressure, stress, and sleep quality in the elderly. Using several databases such as Scopus, Proquest, PubMed, and Google Scholar. Then, the found articles were checked for duplication using Mendeley, resulting in 327 articles. After the title screening process, 129 articles remained. Then 25 articles were included and 15 articles were excluded because they did not meet the study quality.

**Results:** From the review of 10 journals, two main topics were identified, including the effect of progressive muscle relaxation on lowering blood pressure, reducing stress and anxiety, stabilizing vital signs, and improving sleep quality, and the effect of slow stroke back massage therapy on lowering blood pressure, reducing stress and anxiety, and improving sleep quality.

**Conclusion:** Progressive muscle relaxation and slow stroke back massage therapy have a significant impact on lowering blood pressure, reducing stress, and improving the sleep quality of the elderly.

## Introduction

In this era of globalization. The world’s population growth is increasing. With the high birth rates in developing countries, the demand for healthcare services is also increasing. With the increase in growth, it is hoped that everyone will also have a high level of health, considering the potential and risks that can occur to each individual due to unhealthy lifestyles. This triggers several conditions in individuals at risk of diseases such as hypertension, diabetes, and other chronic illnesses. The growth of the elderly population is currently experiencing a significant increase in the 21st century, with the elderly population reaching 465 million in 2000. And it is estimated to increase twofold in 2025 (Winda Yuniarti & Umu Hani Edi Nawangsih, 2017). Aging is a physiological process that occurs in humans, as evidenced by the decline in physiological functions and the human body’s immune system. The aging process is a collection of progressive symptoms that occur due to the decline in human physical condition, which carries the risk of diseases resulting from the decreased function of organs in the human body. Diseases commonly encountered in the elderly include several conditions related to blood circulation system disorders, hormonal metabolism system disorders, and various bone and joint neoplasms (Kusumoningtyas & Ratnawati, 2018).

Progressive muscle relaxation is a systematic technique used to achieve deep states of relaxation and has been proven to improve the quality of life related to both medical and non-medical conditions. The goal is to consciously experience the difference between tension and relaxation, similar to applying pressure to the muscles, where all the muscles in the body, from the tips of the head to the tips of the toes, which were initially tense, become relaxed after receiving progressive pressure (Hendra Irawan et al., 2024).

Based on the research results by (Abdelaziz et al., 2024), preoperative relaxation techniques such as deep breathing relaxation and progressive muscle relaxation significantly provide relaxation effects, reduce heart rate and blood pressure, and are able to reduce pain scales compared to the control group that was not given anything. The combination of relaxation techniques and the administration of ketamine is very effective in reducing pain and lowering manual and hemodynamic feelings post-operation. This is in line with the research conducted by (Pathan et al., 2023), which explains that the combined intervention of slow breathing exercises and progressive muscle relaxation is more effective in reducing heart rate, respiratory rate, blood pressure, and anxiety measured by the perceived stress scale in patients with essential hypertension compared to the intervention alone.

Based on the background and preliminary results that have been explained, the researcher is interested in discussing the influence of modified progressive muscle relaxation slow stroke back massage on reducing blood pressure and stress and improving sleep quality in the elderly. In the background, it is explained that progressive muscle relaxation and slow stroke back massage are among the non-pharmacological therapies that are effective in lowering blood pressure, reducing stress, and improving sleep quality in the elderly. It is hoped that through this therapy, it can have a more effective impact in lowering blood pressure, reducing stress, and improving sleep quality in the elderly.

## Method

In this study, the review framework is based on the guidelines developed by Arksey & O’Malley (2005) and Danielle Levac & Heather Colquhoun (2010), which is carried out using five stages: identifying the research question, identifying relevant studies and then selecting studies, mapping the data, synthesizing and summarizing, and reporting the results. The main objective of conducting this scoping review is to map the available empirical evidence on the topic of interest and identify gaps in the literature. The strength of the scoping review methodology lies in its ability to provide meaning and significance to the subject of interest and to extract the essence from a diverse body of evidence. This scoping review is used to examine the coverage, scope, and nature of relevant research (Davis et al., 2009). Reporting on the search follows the PRISMA flow diagram recommendations (Page et al., 2021).

### Literature search

The research was conducted through a literature search using the Scopus, ProQuest, PubMed, and Google Scholar databases from 2019 to 2024. The literature search used the keywords “progressive muscle relaxation,” “slow stroke back massage,” “blood pressure,” “stress,” “sleep quality,” and “hypertension.” The literature search yielded 1,157 articles filtered using the above keywords, with the following breakdown: Scopus 249 articles, ProQuest 438 articles, PubMed 449 articles, and Google Scholar 21 articles. Then, a feasibility selection was conducted with the full text, resulting in 31 articles for the next selection, of which 25 articles were included and 15 articles were excluded due to not meeting the study quality.

### Data collection

The data collection process was carried out by identifying literature in several databases, including Scopus, Proquest, PubMed, and Google Scholar. Using the PICOS keywords as follows:

P : The population consists only of adults and the elderly.

I : The intervention discusses the problem, focusing on progressive muscle relaxation and slow stroke back massage

C : -

O : stress, sleep quality, and blood pressure

S : randomized control trial, quasy experiment, cross sectional, qualitative

From the literature search results, 1157 articles were found in each database, including Scopus with 249 articles, Proquest with 438 articles, PubMed with 449 articles, and Google Scholar with 21 articles. Then, the found articles were checked for duplication using Mendeley, resulting in 327 articles. After the title screening process, 129 articles remained. Then, a feasibility selection with full texts was conducted, resulting in 31 articles for the next selection, of which 25 articles were included and 15 articles were excluded due to not meeting the study quality.

Two reviewers independently review each publication using the template described below. Not all fields apply to all reviewed publications. At each stage of the review process, two reviewers independently review and make assessments. Covidence indicates discrepancies in the assessments, and a third author reviews alongside the two initial authors who complete the review. The selection of publications can be seen in the PRISMA flow diagram in Figure 1.

**Figure 1.**
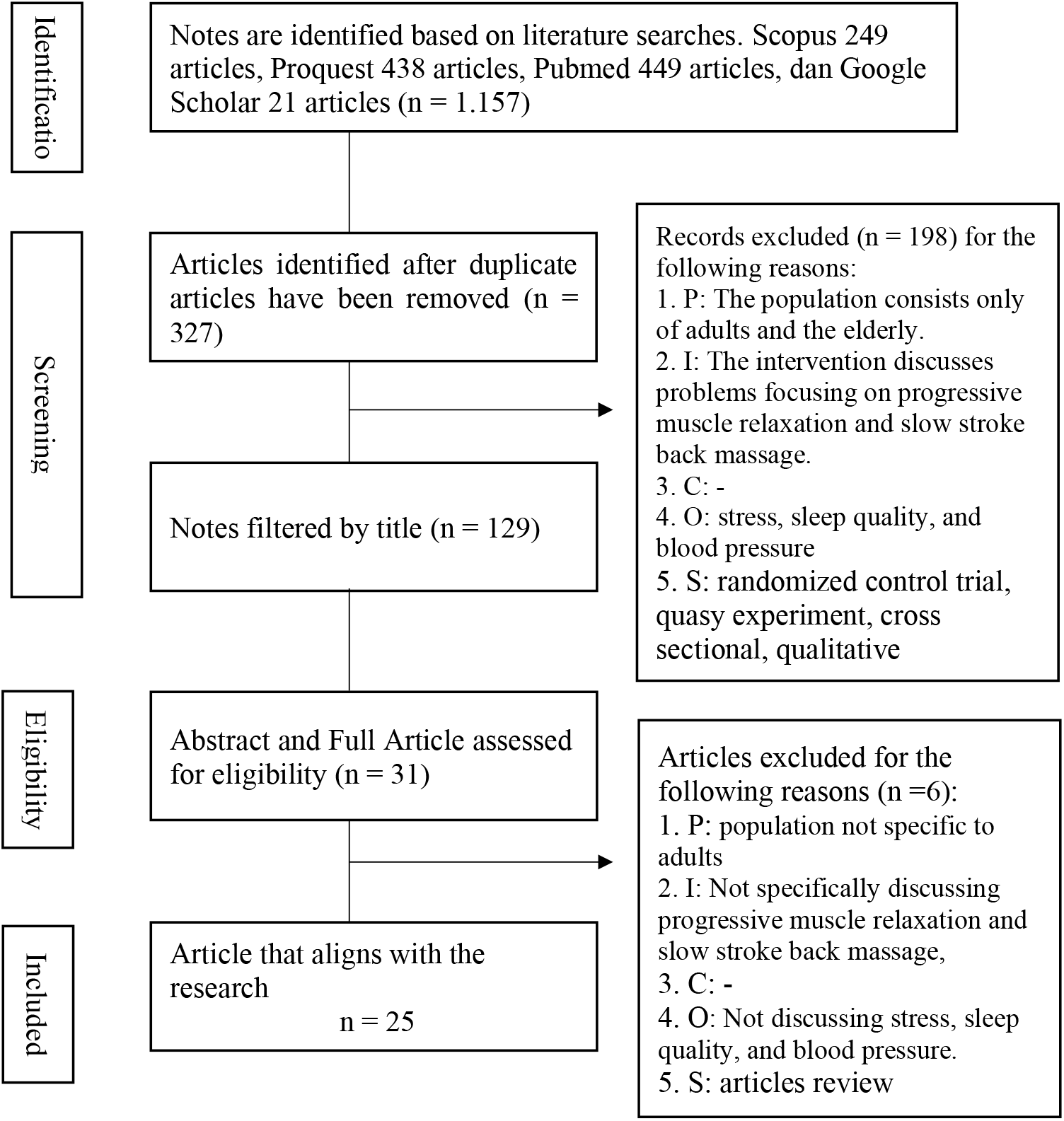
Flow diagram Literature Search Based on PRISMA for the review: The Effect of Modified Slow Stroke Back Massage Progressive Muscle Relaxation on Blood Pressure, Stress, and Sleep Quality in the Elderly.

### Data Extraction and Analysis

For the process of standardizing data extraction procedures, a confidence table was created. Table 1 displays the extraction from 10 literature studies containing the following information: title, author, year, study objective, research design, participants, sample size, variables, outcomes, and research results. Data was extracted and exported from Convidance into a Microsoft Excel spreadsheet for further analysis to explore answers to the stated research questions using qualitative content analysis (Mayring, 2014). The result of developing the inductive theme is the logical categorization of information from the included research based on the principles of thematic analysis. The methodological process consists of eight steps, including the formulation of research questions and the description of the theoretical background, the definition of themes used as selection criteria to determine relevant material from the subject of analysis, and text coding, including the formulation of themes and the assigned research text. Next, the analysis focuses on revising categories/themes and comparing them with the research questions, final coding, developing main themes, and presenting summative and narrative results.

| No | Author/Year /Title | Research Objectives | Study Design | Sample size and quantity | The output that measured | Research result |
| --- | --- | --- | --- | --- | --- | --- |
| 1. | (Pathan et al., 2023)<br>Effect of slow breathing exercise and progressive muscle relaxation technique in the individual with essential hypertension<br>A randomized controlled trial. | to determine the effect of a combination of slow breathing exercises and progressive muscle relaxation techniques on blood pressure, heart rate (HR), respiration rate (RR), and anxiety in patients. diagnosed with essential hypertension. | Quantitative Research ( RCT ) | 64 patients with essential hypertension were divided into 4 sample groups. | 1. Systolic blood pressure<br>2. Diastolic blood pressure<br>3. Heart rate<br>4. Respiratori rate<br>5. Stress. | The main research finding is that the combined intervention of slow breathing exercises and progressive muscle relaxation is more effective in reducing heart rate, respiratory rate, blood pressure, and anxiety (measured by the perceived stress scale) in patients. with essential hypertension compared to the intervention alone. |
| 2. | (Azmy et al., 2021)<br>Efficacy of sport and slow stroke back massage as a non-pharmacolog ical therapy | The objective of this research is to determine the efficacy and effectiveness of sport massage and slow stroke back massage as non-pharmacological therapies to lower blood pressure in elderly individuals with hypertension. | Quantitative Research ( RCT ) | 24 participants in each of the 3 groups (sports massage, slow back massage, control) | The main outcomes measured in this study are changes in systolic and diastolic blood pressure before and after the intervention of sport massage and slow stroke back massage. | 1. Sports massage and slow back massage are both effective in lowering systolic and diastolic blood pressure in elderly patients with hypertension.<br>2. Sports massage is more effective than slow back massage in lowering systolic and diastolic blood pressure.<br>3. Both massage interventions were more effective than the control group in lowering blood pressure. |
| 3. | (Gökşin & Aşiret, 2021)<br>The effect of progressive muscle relaxation on the adaptation of elderly women to | to evaluate the effect of progressive muscle relaxation (PMR) exercises on the level of depression in elderly women and their adaptation to old age. | Quantitative Research ( RCT ) | 49 people and divided into 2 groups. The Intervention Group consisted of 21 people and the Control Group consisted of 28 people. | 1. The effect of progressive muscle relaxation on the level of depression in elderly women<br>2. The influence of progressive | 1. The intervention group that received progressive muscle relaxation (PMR) training for 8 weeks showed a statistically significant improvement in depression levels and adaptation to aging |
|  | depression and old age: a randomised clinical trial |  |  |  | muscle relaxation on the adaptation of elderly women | compared to the control group.<br>2. The adaptation of the intervention group to aging (measured by the ASADE scale) significantly increased after the PMR intervention, while the control group did not show significant changes.<br>3. The depression levels of the intervention group (measured by the GDS-15 scale) significantly decreased after the PMR intervention, while the control group did not show any significant changes. |
| 4. | (Ismansyah et al., 2019)<br>The Effect of Progressive Muscle Relaxation and Slow Deep Breathing Toward Vital Signs of Patients' Hypertension in The Working Area of Bengkuring Medical Center | This study aims to determine the effect of progressive muscle relaxation and slow breathing on the vital signs (blood pressure, pulse, respiration, and body temperature) of hypertensive patients. | Quantitative Research (Quasi Experiment ) | 22 respondents, with 11 respondents each from two treatment groups (progressive muscle relaxation and slow deep breathing) | The outcomes measured in this study are the vital signs of hypertensive patients, which include systolic blood pressure, diastolic blood pressure, pulse rate, respiration, and body temperature. | 1. Progressive muscle relaxation has a significant effect on the vital signs of hypertensive patients, including systolic blood pressure, diastolic blood pressure, pulse rate, respiration, and body temperature.<br>2. Slow deep breathing also has a significant impact on the vital signs of hypertensive patients, including systolic blood pressure, diastolic blood pressure, pulse rate, respiration, and body temperature.<br>3. There is a significant difference in the effects of progressive muscle relaxation and slow deep breathing on systolic blood pressure, pulse rate, and respiration, but not on diastolic blood pressure and body temperature. |
| 5. | (Gultom & Sinaga, 2024) | The objective of this research is to examine the | Qualitative research | The subject in this case study uses one of the | The main outcome measured in this | The research results show that progressive muscle relaxation therapy is |
|  | The Application of Progressive Muscle Relaxation Therapy on Chronic Pain in the Elderly with Hypertension: A Case Study | application and effectiveness of progressive muscle relaxation therapy in the treatment of chronic pain and the improvement of sleep patterns in elderly individuals with hypertension. | (Case Study) | citizens adjusted to the criteria. | study is the effectiveness of progressive muscle relaxation therapy in reducing chronic pain and improving sleep patterns in elderly patients with hypertension. | effective in reducing chronic pain and lowering blood pressure in elderly patients with hypertension. |
| 6. | (Abed Atiyah & Majeed Hameed, 2024)<br>Effect of Relaxation Techniques on Hypertensive Patients' Physiological Outcomes | The objective of this research is to determine the effect of relaxation techniques (Progressive Muscle Relaxation and Deep Breathing) on the physiological parameters of hypertensive patients (systolic and diastolic blood pressure, respiratory rate, heart rate, and oxygen saturation). | Quantitative Research (Quasi Experiment ) | Purposive (non-probability) sampling of 100 hypertensive patients, divided into 50 in the study group and 50 in the control group. | The main outcomes measured in this study are the effects of relaxation techniques (progressive muscle relaxation and deep breathing) on the physiological parameters of hypertensive patients, including systolic and diastolic blood pressure, respiratory rate, heart rate, and oxygen saturation. | 1. The research found a significant decrease in systolic and diastolic blood pressure, heart rate, pulse, and oxygen saturation in the intervention group that received relaxation techniques (progressive muscle relaxation and slow deep breathing), compared to the control group.<br>2. The researchers concluded that slow deep breathing and progressive muscle relaxation can be effective in lowering blood pressure and other physiological actions in hypertensive patients.<br>3. The researchers recommend combining pharmacological therapy with non-drug relaxation technique interventions to manage hypertension and improve |

|  |  |  |  |  |  | physiological parameters in patients. |
| --- | --- | --- | --- | --- | --- | --- |
| 7. | (Patonengan et al., 2023)<br>Effectiveness of Slow Stroke Back Massage (SSBM) on blood pressure, anxiety, and depression among older people with hypertension : A quasi-experimental study | The objective of the research is to determine the effectiveness of slow-motion back massage (SSBM) in reducing blood pressure, anxiety, and depression in elderly patients with hypertension. | Quantitative Research (Quasi Experiment ) | 50 samples divided into 2 groups. The experimental group consists of 25 people and the control group consists of 25 people. | The main research findings of this study are that Slow Stroke Back Massage (SSBM) is effective in reducing blood pressure, anxiety, and depression in elderly patients with hypertension, compared to the control group that received standard therapy. | The main research finding is that the SSBM intervention is effective in significantly reducing blood pressure, anxiety, and depression compared to the control group. The SSBM intervention group showed a strong effect in lowering blood pressure, a moderate effect in reducing anxiety, and a low effect in reducing depression. |
| 8. | (Abdel-Wahab Afifi Araby Ali et al., 2024)<br>Effect of Progressive Muscle Relaxation Techniques on Physiological Parameters, Psychological Factors and Sleep Quality among Pregnant Women with Preeclampsia | The aim of the research is to evaluate the effect of progressive muscle relaxation techniques on physiological parameters, psychological factors (anxiety and stress), and sleep quality in preeclamptic pregnant women. | Quantitative Research (Quasi Experiment ) | Purposive sample of 60 pregnant women with mild preeclampsia, 30 in each group | 1. Physiological parameters: Blood pressure (systolic and diastolic), respiratory rate, and pulse rate<br>2. Psychological factors: Levels of anxiety and stress.<br>3. Sleep quality | 1. The intervention of progressive muscle relaxation techniques caused a significant improvement in the study group compared to the control group in terms of knowledge about preeclampsia, sleep quality, physiological parameters such as blood pressure and respiratory rate, and levels of anxiety and stress<br>2. The total scores of knowledge and sleep quality in the study group showed a significant increase post-intervention and two weeks post-intervention compared to pre-intervention, whereas the control group showed minimal improvement.<br>3. Physiological parameters, total anxiety levels, and total stress levels significantly decreased in the study group post-intervention and two weeks post-intervention compared to the control group. |
| 9. | (Shintia et al., 2023)<br>Combined Murrotal Therapy And Slow Stroke Back Massage (Timun Mas)<br>Lowered Blood Pressure In The Elderly: A Case Study | The objective of the research in this paper is to determine the effect of the combination of murottal therapy and slow stroke back massage (TiMun Mas) on lowering blood pressure in the elderly. | Quantitative Research ( Case study ) | 3 People | The objective of the research in this paper is to determine the effect of the combination of murottal therapy and slow stroke back massage (TiMun Mas) on lowering blood pressure in the elderly. | The research results indicate that the combination of murottal therapy and slow stroke back massage (TiMun Mas) significantly lowers blood pressure in the elderly, as evidenced by a statistically significant decrease in systolic and diastolic blood pressure after the intervention. |
| 10. | (Hsu et al., 2019)<br>Back massage intervention for improving health and sleep quality among intensive care unit patients | The objective of this study This study examines the effects of back massage on improving vital signs, sleep quality, anxiety, and depression among ICU patients. | Quantitative Research (Quasi Experiment ) | Experimental sample (n=30) and control (n=30) | cardiovascular parameters (heart rate, systolic blood pressure, and diastolic blood pressure) and heart rate variability (HRV) parameters (Root Mean Square Of Successive Differences (RMSSD)), High Frequency (HF) power, and Low Frequency (LF) power. | 1. Back massage improves sleep quality in ICU patients.<br>2. Significant reduction in anxiety was observed on the third day.<br>3. No significant effects on heart rate or mean arterial pressure.<br>4. Back massage increases total sleep time and sleep efficiency.<br>5. Subjective and objective sleep measures improved after back massage |

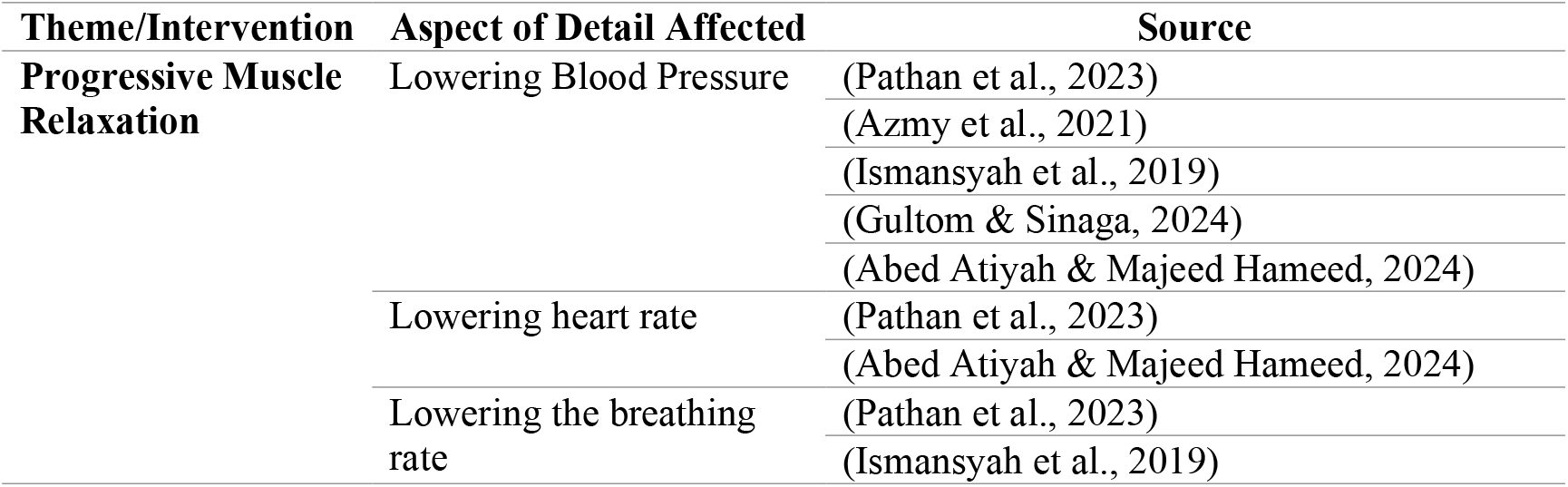
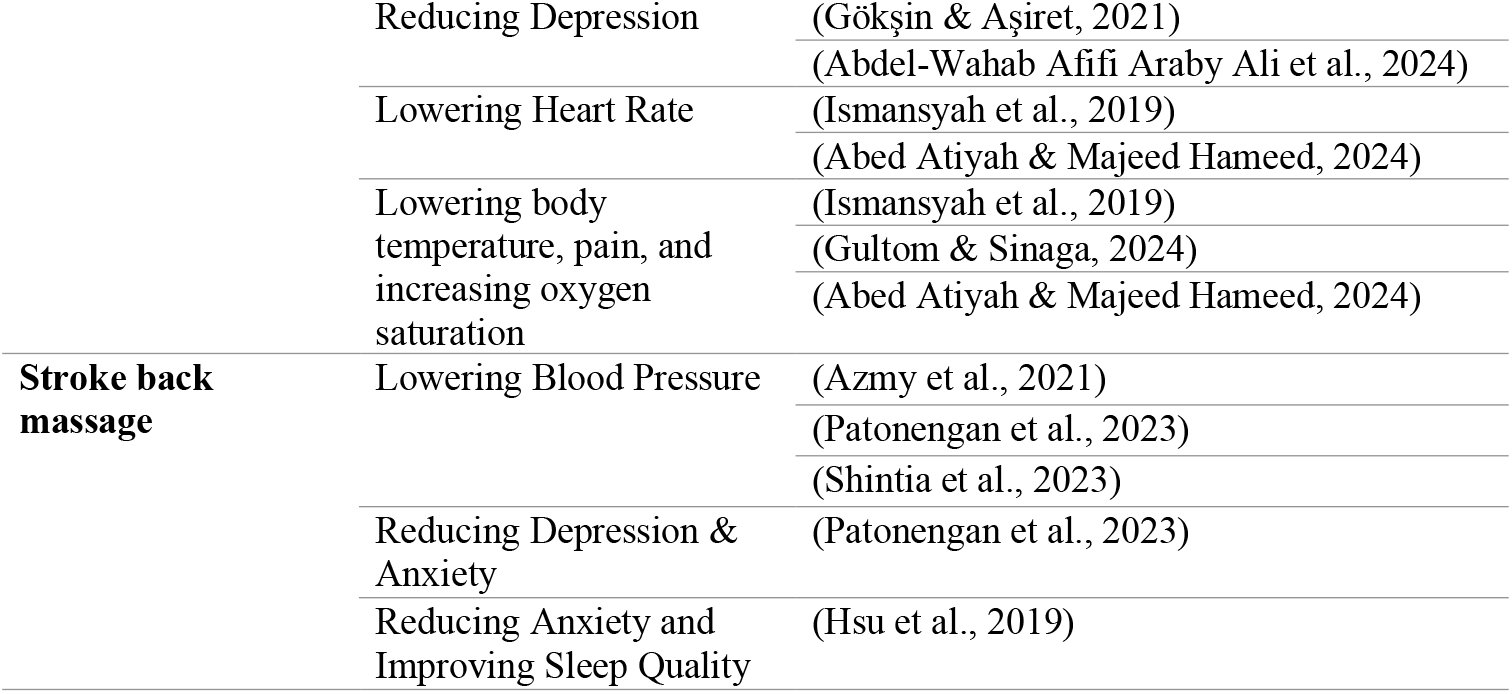

## Discussion

Based on the findings above, there are two discussions that have an influence between progressive muscle relaxation and stroke back massage. In the study, it was determined that progressive muscle relaxation has an effect on lowering blood pressure through 5 studies conducted by (Pathan et al., 2023)., (Azmy et al., 2021)., (Ismansyah et al., 2019)., (Gultom & Sinaga, 2024)., and (Abed Atiyah & Majeed Hameed, 2024). Next, the relationship between muscle relaxation and heart rate reduction was obtained from two studies conducted by (Pathan et al., 2023) & (Abed Atiyah & Majeed Hameed, 2024). Reducing the respiratory rate is derived from (Pathan et al., 2023) and (Ismansyah et al., 2019). Reducing depression resulting from the research findings of (Gökşin & Aşiret, 2021) and (Abed Atiyah & Majeed Hameed, 2024) and lowering body temperature, reducing pain, and increasing oxygen saturation are produced from three studies, namely (Ismansyah et al., 2019), (Gultom & Sinaga, 2024) and (Abed Atiyah & Majeed Hameed, 2024). Then, in the second discussion, the relationship between slow stroke back massage has an effect on lowering blood pressure from three studies presented by (Azmy et al., 2021), (Patonengan et al., 2023), dan (Shintia et al., 2023), reducing depression and anxiety from research (Patonengan et al., 2023), and improving sleep quality based on research findings (Hsu et al., 2019).

From the research results of (Pathan et al., 2023), (Azmy et al., 2021), (Ismansyah et al., 2019), (Gultom & Sinaga, 2024) and (Abed Atiyah & Majeed Hameed, 2024) explained the process of blood pressure reduction in respondents who were given progressive muscle relaxation, this intervention shows a significant effect that the provision of muscle relaxation can help the process of lowering blood pressure if given gradually. This is supported by the research conducted by (Pathan et al., 2023) and (Abed Atiyah & Majeed Hameed, 2024), that muscle relaxation can lower heart rate, physiologically a relaxed heart rate will affect blood pressure. Similarly, it also lowers the pulse, body temperature, pain, and increases oxygen saturation. Slow Stroke Back Massage also has a significant influence on the process of lowering blood pressure as explained by the research findings (Azmy et al., 2021), (Patonengan et al., 2023) and (Shintia et al., 2023). *Slow Stroke Back Massage* intervention can also help reduce depression, anxiety, and improve sleep quality.

### The Effect of Progressive Muscle Relaxation on Lowering Blood Pressure, Reducing Stress, and Improving Sleep Quality in the Elderly

Various impacts influenced by the intervention of progressive muscle relaxation include lowering blood pressure, stabilizing vital signs, and reducing excessive stress and anxiety. (Abed Atiyah & Majeed Hameed, 2024). Muscle relaxation is performed by consciously contracting and stretching the muscles. When the muscles contract, the nerve impulse reaches the axon terminal, causing the release of acetylcholine, which will diffuse across the synapse. Acetylcholine will make the sarcolemma more permeable to Na+ ions, which will quickly enter the cell. The sarcolemma undergoes depolarization. The depolarization process will stimulate the release of Ca2+ from the sarcoplasmic reticulum. The Ca2+ ions will bind to the troponin-tropomyosin complex, causing the actin filaments to shift further apart. Myosin will break down ATP to release its energy, the bridge on the myosin will then attach to the actin filament and pull it towards the center of the sarcomere, causing the sarcomere to shorten. The entire sarcomere in the muscle fibers will shorten, causing contraction in the entire muscle fibers. When the sarcolemma repolarizes, K+ ions leave the cell, restoring the positive charge outside the cell and the negative charge inside the cell. This pump will then return Na+ ions out of the cell and K+ ions into the cell, while cholinesterase in the sarcolemma will deactivate acetylcholine. Muscle fibers will relax and return to their original length. (Endar Sulis Tyani & N, 2015).

Progressive muscle relaxation will trigger a decrease in heart pumping activity and cause the arteries to dilate, resulting in a significant amount of fluid leaving the blood circulation. This will reduce the workload of the heart because hypertensive patients have a faster heart rate to pump blood due to increased blood pressure. (Azizah, 2015). The process of relaxation of heart pumping activity will decrease, arteries will dilate, and a lot of fluid will exit the circulation. The walls of elastic and easily distensible arterial blood vessels will easily expand the diameter of the blood vessel walls to accommodate changes in pressure. The ability of arterial distension prevents fluctuations in blood pressure. (Endar Sulis Tyani & N, 2015). This is proven by research conducted by (Pathan et al., 2023) (Azmy et al., 2021), (Ismansyah et al., 2019) (Gultom & Sinaga, 2024) and (Abed Atiyah & Majeed Hameed, 2024). That the intervention carried out by providing progressive muscle relaxation significantly lowers blood pressure.

In addition, progressive muscle relaxation also has an effect on anxiety conditions and reduces stress levels. Based on the research conducted by (Gökşin & Aşiret, 2021) and (Abed Atiyah & Majeed Hameed, 2024) that, the provision of progressive muscle relaxation can reduce levels of anxiety and depression. This is because progressive muscle relaxation triggers the parasympathetic nervous system. The parasympathetic nervous system triggers the release of endorphins in the body, which have a calming and soothing effect. At certain times, this will suppress the release of cortisol and adrenocorticotropic hormone, resulting in a decrease in anxiety scores. Moreover, in such a state, it will improve psychological and physiological conditions and suppress disturbances in the central and autonomic nervous systems. (Abdel-Wahab Afifi Araby Ali et al., 2024).

The influence of Slow Stroke Back Massage on lowering blood pressure, reducing stress, and improving sleep quality.

The provision of Slow Stroke Back Massage has an impact on lowering blood pressure, reducing depression and anxiety, and improving sleep quality. Based on the research conducted by (Azmy et al., 2021), and two other studies that the intervention provided through Slow Stroke Back Massage has an effect on lowering blood pressure in hypertensive patients. In the study (Patonengan et al., 2023) and (Hsu et al., 2019) also explains that Slow Stroke Back Massage reduces depression and lowers anxiety in patients. A back massage with slow movements on the back and shoulders, applying pressure on the skin with gentle strokes, provides pleasure and comfort, while short and circular strokes stimulate the muscles. The administration of slow stroke back massage therapy will lower blood pressure. The advantages of this therapy include that relaxation therapy is easier to perform, safe for the elderly, simple, and inexpensive. (Yuliyanto & Abdurrachman, 2021). The Slow Stroke Back Massage therapy is a massage applied to the skin with pressure on the back area using stroking, petrissage, and friction techniques. This massage provides nerve stimulation to the superficial skin, which is then transmitted to the brain’s hypothalamus, triggering the release of endorphins. Endorphins provide a relaxation effect that results in vasodilation of blood vessels, thereby lowering blood pressure. (Setiawan et al., 2023).

### Implication

The provision of relaxation techniques such as progressive muscle relaxation and slow stroke back massage is a good technique to be performed in the nursing intervention process for elderly patients. However, in the process of administering it, the physiological and psychological conditions of the elderly must be taken into account, and it is necessary to identify the physical condition regarding injuries, wounds, or the mobility ability of the elderly. Several studies have explained the results of administering progressive muscle relaxation and slow stroke back massage, which are effective in lowering blood pressure, reducing depression and anxiety, and improving the sleep quality of the elderly. But contraindications were not explained in the study.

This research highlights the importance of therapy provided to the elderly, by examining its effectiveness and impact on several factors resulting from the therapy. Furthermore, the enhancement of the capacity and skills of healthcare workers, especially nurses, needs to be carried out to ensure that the interventions are performed correctly and according to procedures, thereby minimizing errors in providing interventions such as progressive muscle relaxation or slow stroke back massage.

### Limitations

The review of this publication process is quite limited, as most of the references used are international sources where there are differences in the conditions of the elderly in Indonesia. Cultural characteristics, physical conditions, and the provision of healthcare services tend to have quite significant differences. Thus, the explanations from the publication findings cannot be generalized for implementation in specific service locations. There is a need for studies on transcultural, bio-psychosocial aspects and the characteristics of the elderly in various parts of the world. Moreover, the findings of the published journals are mostly quantitative, lacking in-depth statements from the practitioners or the elderly who received the intervention. There is a need for publications that include qualitative research designs to delve deeper into the effectiveness of the therapy provided.

## Conclution

Several studies suggest that progressive muscle relaxation can help lower blood pressure, relieve pain, stabilize vital signs such as breathing, pulse, and heart rate, reduce the risk of depression and anxiety, and improve sleep quality. Similarly, the administration of slow stroke back massage therapy helps lower blood pressure, reduce the risk of depression and anxiety, and improve sleep quality. From the results of the research, progressive muscle relaxation and slow stroke back massage therapy have a significant impact on lowering blood pressure, reducing stress, and improving the sleep quality of the elderly.

## Data Availability

All data produced in the present work are contained in the manuscript

## Funding Source

This research did not receive funding from any institution, commercial or non-profit.

## Conflict of Interest Declaration

The authors declare that they have no financial interests or competing personal relationships that could influence the work reported in this research.

## Notes

### Competing Interest Statement

The authors have declared no competing interest.

### Funding Statement

This study did not receive any funding

